# Consequences of COVID-19 vaccine allocation inequity in Chicago

**DOI:** 10.1101/2021.09.22.21263984

**Authors:** Sharon Zeng, Kenley M. Pelzer, Robert D. Gibbons, Monica E. Peek, William F. Parker

**Author notes:** **Corresponding Author:** William F. Parker, MD, Ph.D., 5841 S Maryland Avenue, MC 6076, Chicago, IL 60637.

## Abstract

During Chicago’s initial COVID-19 vaccine rollout, the city disproportionately allocated vaccines to zip codes with high incomes and predominantly White populations. However, the impact of this inequitable distribution on COVID-19 outcomes is unknown. This observational study determined the association between zip-code level vaccination rate and COVID-19 mortality in residents of 52 Chicago zip codes. After controlling for age distribution and recovery from infection, a 10% higher vaccination rate by March 28, 2021, was associated with a 39% lower relative risk of death during the peak of the spring wave of COVID-19. Using a difference-in-difference analysis, Chicago could have prevented an estimated 72% of deaths in the least vaccinated quartile of the city (vaccination rates of 17.8 – 26.9%) if it had had the same vaccination rate as the most vaccinated quartile (39.9 – 49.3%). Inequitable vaccine allocation in Chicago likely exacerbated existing racial disparities in COVID-19 mortality.

## INTRODUCTION

As of September 2021, the COVID-19 pandemic has hospitalized nearly 30,000 and killed over 5,800 people in Chicago.^1^ Black and Latinx residents in zip codes with the lowest incomes have been disproportionately affected by COVID-19, accounting for over 70% of COVID-19 deaths despite making up 60% of Chicago’s population.^2,3^ Previous studies have found a spatial correlation between the percentage of residents identifying as Black within a Chicago community area and COVID-19 death rates, comorbidities, and social vulnerability.^4^ After the FDA approved novel COVID-19 vaccines with an Emergency Use Authorization in December 2020, a common goal of phased implementation plans across the country was to mitigate racial inequities in COVID-19 outcomes. However, White and high-income communities were often the first people to receive vaccines in major cities.^5^ Structural racism, defined as the differential access to goods, services, and opportunities by race, has played a substantial role in COVID-19 disparities. For example, in Chicago, structural racism manifested in part through online-only vaccine signups and geographic concentration of vaccination clinics in affluent areas; low-income racial/ethnic minority populations had disproportionately less access during initial phases of vaccine distribution.^6^ However, the consequences of the inequitable vaccine allocation are unknown.

The Chicago Department of Public Health provides the public some of the most granular COVID-19 data in the county, releasing weekly updates in cases, deaths, and vaccinations by resident zip code. In this study, we use this detailed geospatial data to quantify the association between vaccinations and COVID-19 mortality in the spring wave of 2021.

## METHOD

### Study Design and Population

This study was a secondary analysis of publicly available, de-identified data and was granted exemption status by the University of Chicago Biological Sciences Division/University of Chicago Medical Center Institutional Review Board. We used datasets from the Chicago Department of Public Health (CDPH)^7,8^ comprising COVID-19 infections, deaths, and vaccinations organized by zip code from August 8, 2020, through June 13, 2021 (last updated September 8, 2021). We excluded zip codes with less than 10,000 residents or were primarily outside city limits. Population counts, demographic data, and socioeconomic data were obtained from the US Census Bureau American Community Survey (ACS) 5-year estimates for 2015-2019.^9,10^ Demographic data in the Census, including race and ethnicity responses, were based upon self-identification.

Because the first dose of a two-dose vaccine series provides significant protection against COVID-19 infection and severe outcomes, the exposure of interest was the 6-week lagged percentage of zip code residents who had received at least one dose of any COVID-19 vaccine.^11–14^ To compare the average effect of vaccinations, we divided the zip codes into groups based on the percentage of residents who had received at least one dose by March 28, 2021. We defined three groups of zip codes (least vaccinated, intermediate, and most vaccinated) representing the bottom quartile, interquartile range, and top quartile of first-dose vaccination rates. This date was six weeks before the peak of the spring wave of deaths and marked the end of severe vaccine scarcity in Chicago.^15^

To investigate the contribution of structural racism to vaccination inequities, we compared demographic data and indicators of socioeconomic status between the vaccination groups. To analyze differences in age, sex, race, and ethnicity between groups, we calculated the total number of people belonging to each category and the population-weighted average percentage of residents in each category. To calculate the high school graduation and the health insurance rates, we followed the US Census Bureau reporting convention and used population denominators of individuals over 25 years old and civilian noninstitutionalized individuals.^9^ To find the median household income of a vaccination group, we calculated the population-weighted median zip code-level household income.

### Main Outcomes and Measures

The primary outcome was deaths from COVID-19, defined by the CDPH as occurring among confirmed cases with a positive molecular (PCR) or antigen test,^7^ in a given zip code during the spring wave of deaths in Chicago (March 28, 2021, to June 13, 2021). The secondary outcome was weekly new infections in each zip code defined by the CDPH as a positive PCR or antigen test.^7^

### Statistical Analysis

We assessed the association of vaccination coverage with COVID-19 mortality with three different approaches. First, we calculated an unadjusted Pearson product-moment correlation coefficient between vaccination level and total spring wave mortality, weighted by zip code population). Using bootstrapped standard errors, we ran a t-test for significance with a null hypothesis of no correlation (r = 0).

Second, we conducted a difference-in-difference analysis to estimate the number of lives that could have been saved in the least vaccinated group, accounting for baseline differences in COVID-19 death rates between the groups. We took the difference from week to week in the most vaccinated group and applied the same trend to the least vaccinated group as a counterfactual for the subsequent spring wave of deaths. The difference between the counterfactual and actual number of deaths provided an estimate of the number of lives that equitable vaccine allocation could have saved in the spring 2021 wave if the least vaccinated group had experienced the same trend in mortality as the most vaccinated group. We performed a test of parallel trends before the spring wave by fitting the data from December 13, 2020, to March 28, 2021, to a Poisson regression model with log link and testing the interaction between vaccination group and decline in death rate **<u>(Table S4)</u>**.

Finally, to estimate the marginal treatment effect of higher vaccination rates on weekly COVID-19 death rates during the spring wave, we fit a mixed-effects Poisson regression model using data from December 13, 2020, to June 13, 2021 **<u>(Table S5)</u>**. Fixed effects in the model were: the number of weeks since the peak of the second wave (December 13, 2020), an indicator variable for the spring wave (after 3/28/21), and the interaction between spring wave and percentage of population vaccinated by March 28, 2021. The interaction between spring wave and vaccination level estimated the average incidence rate ratio of 6-week lagged vaccination levels on weekly mortality during the spring wave. We controlled for the percentage of the population over 65 years old and the percentage recovered from previous COVID-19 infection by March 28, 2021, by including these covariates as fixed effects in the model. Within zip-code correlation of weekly COVID-19 mortality rates was accounted for with a zip code level random intercept, and the offset was the population of each zip code.

### Sensitivity analyses

We performed two robustness checks. First, we changed the exposure to the 6-week lagged percentage of residents fully vaccinated and determined the association with COVID-19 mortality. Next, we re-fit the mixed-effect Poisson regression with vaccination quintile as a categorical variable to assess for non-linear effects of vaccination rate on mortality. **<u>(Supplemental Methods)</u>**.

We performed all analyses with R version 4.0.5 (2021-03-31) and have posted complete datasets and statistical code at https://github.com/zengsharon/ChicagoVaccineAllocation.

## RESULTS

### Vaccination Inequities

There were 52 zip codes included in the analysis, representing an overall population of 2,686,355 Chicago residents **<u>(Table S1)</u>**. We excluded seven zip codes that were mostly outside city limits or had fewer than 10,000 residents **<u>(Table S2)</u>**. Cumulative first dose vaccination rates on March 28, 2021, ranged from 17.8% to 49.3% by zip code of residence **<u>(Figure S1a)</u>**. The first dose vaccination rate ranged from 17.8% to 26.9% in the least vaccinated quartile (13 zip codes, population of 619,518). The intermediate vaccination group (25^th^ – 75^th^ percentile) had vaccination levels ranging from 27.6% to 39.4% (26 zip codes, population 1,582,146). Lastly, the most vaccinated quartile of zip codes had vaccination levels of 39.9% to 49.3% (13 zip codes, total population of 484,691). All of the zip codes which were in the least vaccinated quartile on March 28, 2021 remained the least vaccinated through the spring wave, which peaked six weeks later, while 84.6% (2 out of 13) of the zip codes in the most vaccinated quartile on March 28, 2021 remained in this group through the spring wave **<u>(Figure S1a)</u>**.

### Study Population

Across all 52 zip codes studied, 29% of residents identified as Latinx, 7% identified as Asian Non-Latinx, 29% identified as Black Non-Latinx, and 33% identified as White Non-Latinx **<u>(Table S3)</u>**. Eighty-four percent of residents over 25 years old had at least a high school graduate degree, and 90% of the civilian noninstitutionalized population had health insurance. The median household income was $52,044.

In the least vaccinated quartile, 80% of residents identified as Black Non-Latinx, compared to 8% in the most vaccinated quartile and 16% in the intermediate group. In the intermediate group, 40% of residents identified as Latinx, compared to 11% in the most vaccinated quartile and 13% in the least vaccinated quartile (p < 0.01 for both comparisons). In the most vaccinated quartile, 68% of residents identified as White Non-Latinx and 10% identified as Asian Non-Latinx, compared to 34% and 8% respectively in the intermediate group and 5% and 1% respectively in the least vaccinated quartile. The difference in the distribution of race/ethnicity between groups was statistically significant (p < 0.001). There was no statistically significant difference in distribution of age (p = 0.141) or sex (p = 0.808) between the groups **(Table 1)**.

**Table 1:** Demographics of vaccination groups.

|  | % of Zip Code Population with First Doses by 3/28/21 |  |  | p<sup>b</sup> |
| --- | --- | --- | --- | --- |
|  | [17.8, 26.9] | (28.3,38.8] | (38.8,49.3] |  |
| **Age** |  |  |  |  |
| **No. (%)<sup>a</sup>** |  |  |  |  |
| Age 0-17 | 151766 (24) | 350272 (22) | 59282 (12) | 0.141 |
| Age 18-65 | 374758 (60) | 1046275 (66) | 369739 (76) |  |
| Age 65+ | 92994 (15) | 185599 (12) | 55670 (11) |  |
| **Sex** |  |  |  |  |
| **No. (%)** |  |  |  |  |
| Female | 337097 (54) | 797669 (50) | 243892 (50) | 0.808 |
| Male | 282421 (46) | 784477 (50) | 240799 (50) |  |
| **Race and Ethnicity** |  |  |  |  |
| **No. (%)** |  |  |  |  |
| Latinx | 80974 (13) | 638721 (40) | 54243 (11) | < 0.001 |
| Asian Non-Latinx | 4103 (1) | 121594 (8) | 49523 (10) |  |
| Black Non-Latinx | 498263 (80) | 246648 (16) | 39005 (8) |  |
| White Non-Latinx | 27914 (5) | 538835 (34) | 327806 (68) |  |
| Other Race Non-Latinx | 8264 (1) | 36348 (2) | 14114 (3) |  |
| **High School Graduate or Higher** |  |  |  |  |
| **No. (%)<sup>c</sup>** | 336956 (83) | 883791 (82) | 361664 (95) | < 0.001 |
| **With Health Insurance** |  |  |  |  |
| **No. (%)<sup>d</sup>** | 557180 (91) | 1385479 (89) | 459730 (96) | < 0.001 |
| **Median Household Income** |  |  |  |  |
| **\$ (IQR)** | 34535 (12055) | 53864 (25213) | 94859 (17449) | 0.040 |

<sup>a</sup> No. (%) = total number across the group (percentage of total group population)
<sup>b</sup> Chi-squared tests were used to compare distributions of age (degrees of freedom [df] = 4), sex (df = 2), and race and ethnicity (df = 8) across all three groups. High school graduates and health insurance were compared across all groups using ANOVA (df = 2). Median household income was compared across all groups using a Kruskal-Wallis test (df = 2).
<sup>c</sup> Calculated as percentage of group population over 25 years old
<sup>d</sup> Calculated as percentage of group civilian noninstitutionalized population

The groups also differed significantly in socioeconomic indicators. In the most vaccinated quartile, 95% of residents over 25 years old had a high school graduate degree or higher, compared to 83% in the least vaccinated quartile and 82% in the intermediate group (p < 0.001). Ninety-six percent of noninstitutionalized civilian residents in the most vaccinated quartile had health insurance coverage, compared to 91% in the least vaccinated quartile and 89% in the intermediate group (p < 0.001). The median household income in the most vaccinated quartile was $94,859 [IQR: $17,449], compared to $34,535 [IQR: $12,055] in the least vaccinated group and $53,864 [IQR: $25,213] in the intermediate group (p = 0.040).

### Association of COVID-19 Mortality with Vaccination

The zip codes with the lowest vaccination levels by March 28, 2021 were clustered in the south and west regions of Chicago, primarily consisting of low-income, Black and Latinx communities that experienced the highest spring wave mortality rates **(Figure 1)**. The population-weighted Pearson correlation between vaccination level on March 28, 2021, and deaths per 100,000 across the spring wave was r = −0.77 (p < 0.001).

**Figure 1:**
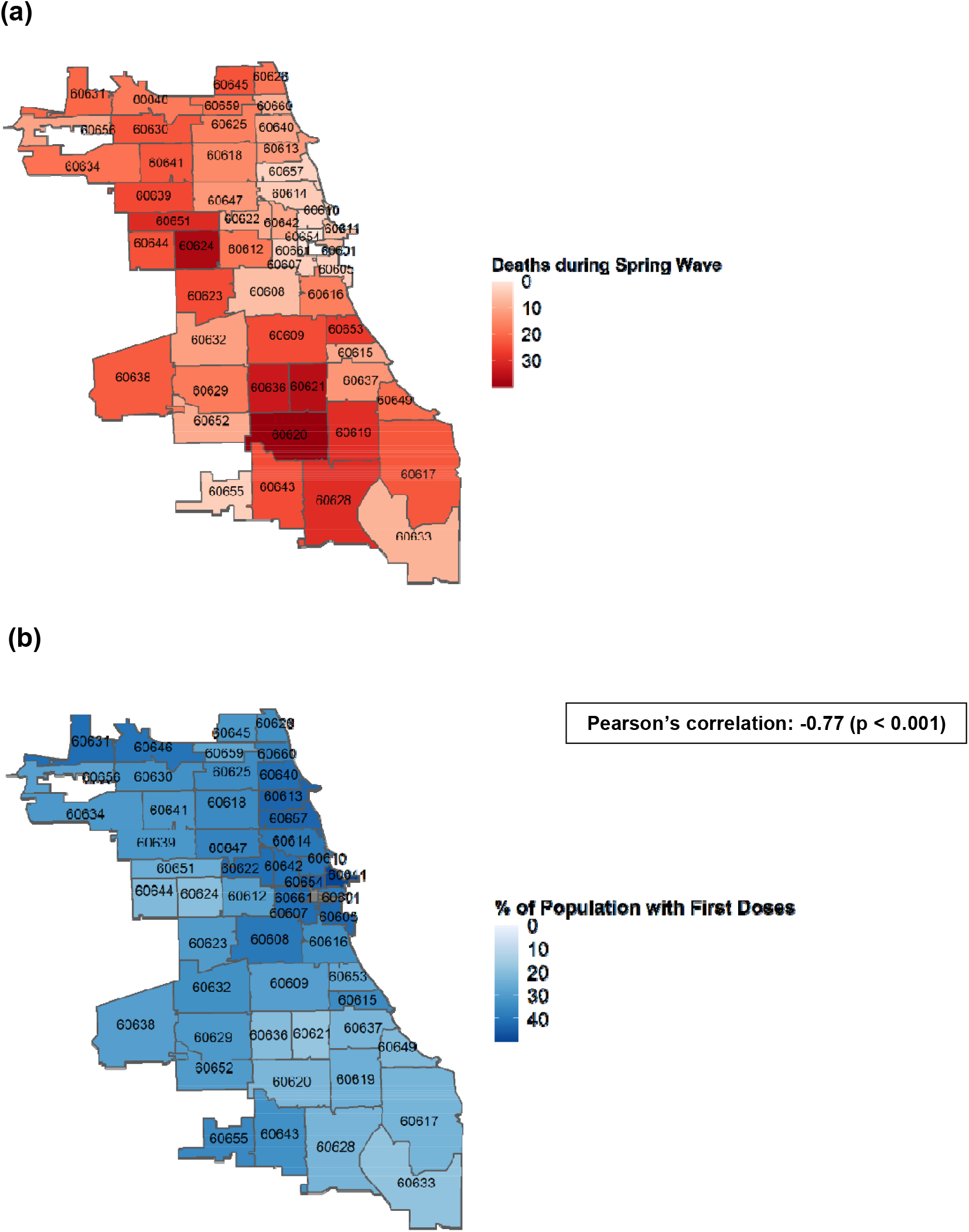
Geographic distribution of vaccinations and spring wave mortality. (**a**) Maps of vaccination levels on March 28, 2021. **(b)** COVID-19 deaths per 100,000 zip code residents from March 28, 2021 to June 13, 2021.

All groups experienced an increase in cases from February 2021 that peaked in April 2021 **(Figure 2a)**. In the spring wave of deaths that followed from March 28, 2021 to June 13, 2021, the least vaccinated and intermediate groups experienced peak rolling average mortality rates of 2.97 and 1.64 deaths per 100,000 people, respectively. In contrast, the most vaccinated group experienced a peak of 0.91 deaths per 100,000 people (**Figure 2b)**.

**Figure 2:**
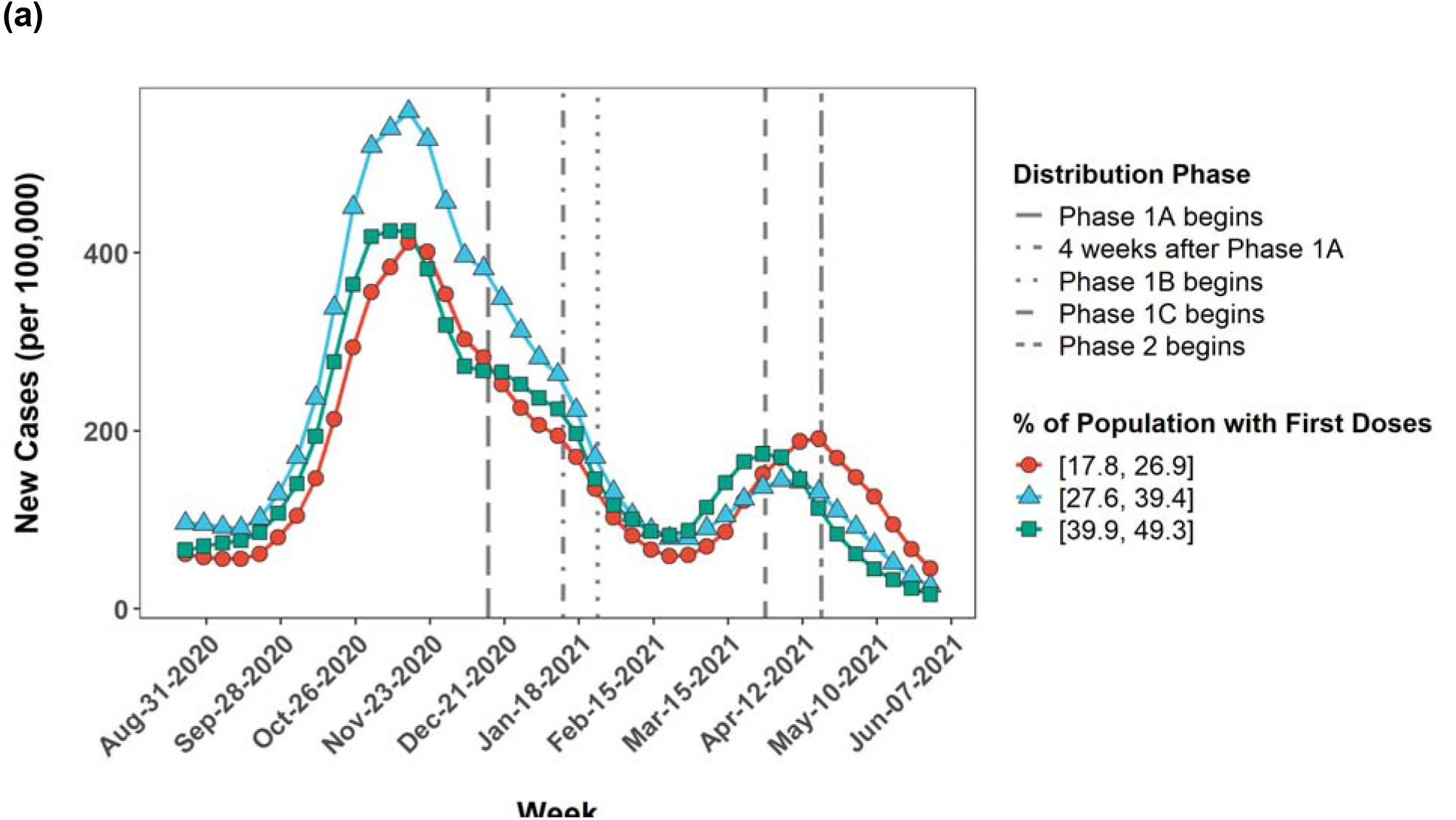
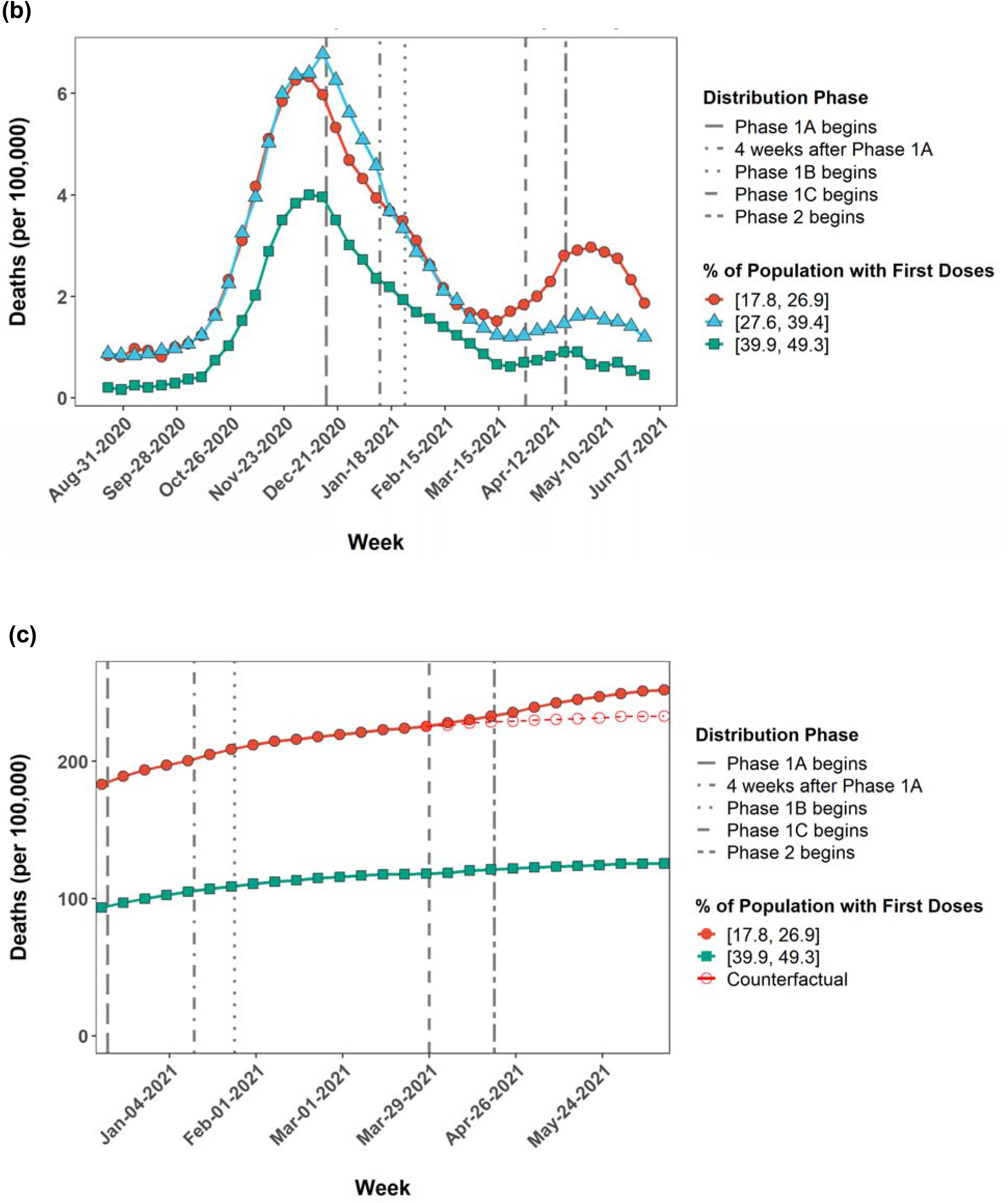
COVID-19 outcomes. **(a)** Line graph of 5-week rolling average of new cases per 100,000 population by vaccination group from August 8, 2020 to June 13, 2021. Centers of the rolling averages are plotted. **(b)** Line graph of 5-week rolling average of deaths per 100,000 population by vaccination group. Centers of the rolling averages are plotted. **(c)** Line graph showing cumulative mortality by vaccination group from December 13, 2020 to June 13, 2021. During the spring wave, there were a total of 164 deaths in the least vaccinated group (filled red circles) compared to 36 deaths in the most vaccinated group (filled green squares). Open red circles indicate the hypothetical scenario in which the least vaccinated group (filled red circles) had experienced the same trend in mortality as the most vaccinated group starting on March 28, 2021. In this scenario, approximately 46 people would have died in the least vaccinated group after accounting for differences in population. Thus, the difference-in-difference estimates that 118 lives could have been saved if the least vaccinated group had levels of vaccination equivalent to the most vaccinated group.

An F-test of Poisson regression analysis confirmed that the vaccination groups experienced parallel trends in weekly mortality during the decline of the second wave of deaths **<u>(Table S4)</u>**. Using difference-in-difference analysis, we estimate that 72% of the deaths in the least vaccinated quartile of the city could have been prevented if the trend in mortality over time had followed most vaccinated quartile (only 46 deaths compared to 164 observed deaths) **(Figure 2c)**.

### The marginal effect of increasing vaccination coverage

In a mixed-effects Poisson regression model, a 10% increase in residents with at least one dose on March 28, 2021 was associated with an incidence rate ratio of 0.61 (CI: 0.51 – 0.72, p < 0.001), corresponding to a 39% decrease in the weekly risk of death from COVID-19. After adjusting for the percentage of residents over 65 years old and the percentage of residents who had recovered from COVID-19 infection **(Table 2)**, the incidence rate ratio was 0.61 (CI: 0.52 – 0.72, p < 0.001). When comparing vaccination groups, the lowest vaccination quartile was associated with a 2.52 times greater weekly risk of death from COVID-19 compared to the highest vaccination quartile **<u>(Table S6)</u>**.

**Table 2:**
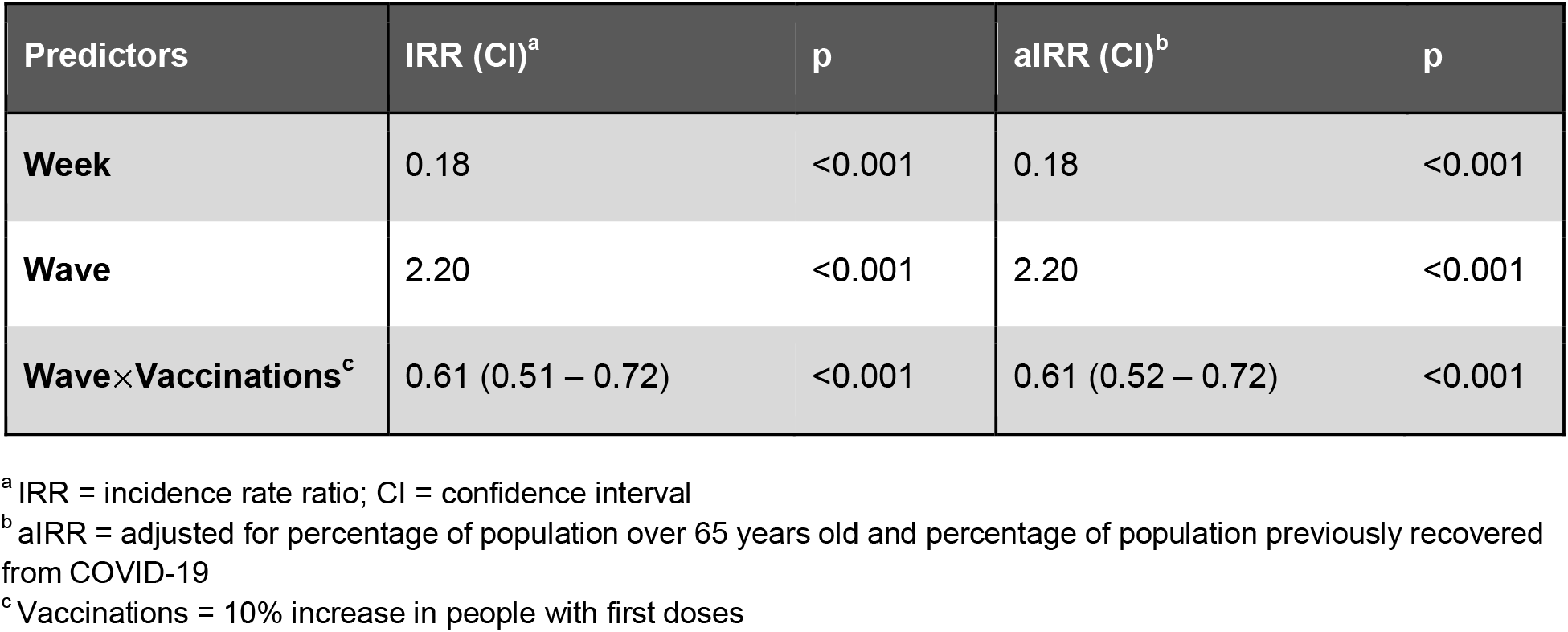
Multivariable analysis of risk reduction in COVID-19 deaths, by vaccinations before spring wave.

In another Poisson regression model examining full vaccination coverage, a 10% increase in fully vaccinated residents on March 28, 2021 was associated with an incidence rate ratio of 0.45, corresponding to a 55% decrease in weekly risk of death from COVID-19 **<u>(Table S6)</u>**.

## DISCUSSION

Early phases of COVID-19 vaccine implementation both in Chicago and nationwide focused on distributing a scarce resource to areas in the most need. We have found that lower vaccination levels by the end of this period of scarcity were negatively correlated with mortality rates during the spring wave of COVID-19 deaths in Chicago. In a mixed-effects Poisson regression analysis, a 10% higher vaccination level was associated with a 39% reduction in the peak of deaths during the spring wave. If the least vaccinated quartile of zip codes had the same vaccination rate as the most vaccinated quartile, the city could have averted a majority of the deaths in these hardest-hit areas. Zip codes that had lower vaccination levels just before the spring wave had more residents who identified as Black and more low-income and uninsured residents, suggesting that structural racism contributed to inequities in vaccine allocation.

This observational study leverages detailed geospatial data to estimate the deadly consequences of vaccine inequity. For the first time in the pandemic, the trends between zip codes were no longer parallel. The highly vaccinated parts of the city were effectively spared from the virus because of priority access to vaccines. Although the most vaccinated group experienced the fewest deaths during the spring wave, this was not reflected in weekly new infections, suggesting that exposure to infection did not account for the differences in mortality. The discrepancy between infections and deaths could indicate that first-dose vaccination levels during our study period were not high enough to sustain herd immunity against infection, or could reflect differences in testing and surveillance practices.

Further research is also needed to clarify and disentangle the mechanisms of vaccine hesitancy and structural racism in vaccine allocation. Vaccine hesitancy and medical distrust in historically marginalized communities, particularly among Americans who identify as Black, partly stems from current and historical structural racism in healthcare.^16,17^ In addition, racial equity in vaccination improved after Chicago put in place a temporary program to vaccinate communities that had disproportionately suffered from severe COVID-19 outcomes, suggesting that problems of access, as opposed to hesitancy, were the primary contributors to unequal vaccination levels in early phases of vaccine allocation.^18^

We focused on the early phases of vaccine implementation as this reflects the period when vaccines were still a scarce resource whose distribution was dependent on allocation policies. Although large academic hospitals had the resources to vaccinate more patients, they were often unreachable for Black and Latinx communities.^19^ Scheduling methods favored people with English fluency and resources to make appointments as soon as web pages refreshed and disadvantaged people of color and immigrants who have less flexibility to take time off work to get vaccinated.^20,21^ Whether through barriers to access, medical distrust, or other factors influencing the decision to pursue vaccination, early vaccine allocation in Chicago failed to provide equitable vaccinations to Black and Latinx communities. This inequality in vaccinations is likely to have played a major role in exacerbating the racial disparity in deaths during the spring wave.

### Limitations

Our study has several limitations. First, we had zip code level (not patient-level) data which limited the study design and our ability to control for observed confounders. Second, unmeasured time-varying zip code level confounders could bias our difference-in-difference estimate. However, the parallel trends between high and low zip codes during the pre-spring wave period make confounding by time-invariant unobserved zip code variables unlikely. Finally, the data’s integrity depends on the accurate recording of the zip code of residence for both vaccinations and deaths in the city of Chicago database. Unhoused persons or people without a permanent address are unlikely to be recorded accurately in the data.

## Conclusions

During Chicago’s initial COVID-19 vaccine rollout, the city disproportionately allocated vaccines to zip codes with high incomes and predominantly White populations. During the following spring 2021 COVID-19 wave, highly vaccinated areas broke with historical mortality trends and had substantially fewer deaths. Inequitable vaccine allocation in Chicago exacerbated existing racial disparities in COVID-19 mortality. Estimates of incidence risk ratios obtained from a mixed-effects Poisson generalized linear model with log link using fixed effects (wave and the relationship between wave and vaccination level) and random effects of zip code on intercept. Adjusted incidence rate ratios were estimated from a second model which included percentage of population over 65 years old and percentage of population previously recovered from COVID-19 infection as fixed effects. For Week and Wave, IRRs are calculated for a zip code with the mean vaccination level on March 28, 2021. Confidence intervals and p-values were obtained from asymptotic Wald tests. Full model output is reported in **<u>Table S5</u>**.

## Data Availability

The data used in this manuscript was is publically available.

https://github.com/zengsharon/ChicagoVaccineAllocation

https://data.cityofchicago.org/browse?limitTo=datasets&sortBy=alpha&tags=covid-19

## ACKNOWLEDGEMENTS

Support was provided by the National Institute on Aging (Grant No. 5T35AG029795-14 to Sharon Zeng as part of the Pritzker Summer Research Program in Aging) and the National Heart, Lung, and Blood Institute (Grant No. K08 HL150291 to William Parker).

