## Supplement for "Consequences of COVID-19 vaccine allocation inequity in Chicago"

Sharon Zeng, BA[1]

Kenley M. Pelzer, PhD[2]

Robert D. Gibbons, PhD[2, 3]

Monica E. Peek MD, MPH, MS[2,4]

William F. Parker, MD, PhD[2,5]

Author affiliations:

[1]Pritzker School of Medicine, University of Chicago, Chicago, Illinois

[2]Department of Medicine, University of Chicago, Chicago, Illinois

[3]Department of Public Health Sciences, University of Chicago, Chicago, Illinois

[4]University of Chicago Division of the Biological Sciences, Chicago, Illinois

[5]MacLean Center for Clinical Medical Ethics, University of Chicago, Chicago, Illinois

**Corresponding Author:**
William F. Parker, MD, Ph.D.
5841 S Maryland Avenue
MC 6076
Chicago, IL 60637


Code used for this project is available at

https://github.com/zengsharon/ChicagoVaccineAllocation

**Figure S1: Visualizations of vaccination groups**

**(a)** Line graph showing cumulative vaccinations in each zip code from December 13, 2020 to June 13, 2021. Color and dot shape represent vaccination groups to which the zip codes were assigned. Square brackets indicate which end points were included in a vaccination group. Vertical lines denote the first date of vaccination phases in Chicago (Phase 1A: residential healthcare facilities and healthcare workers; 4 weeks after Phase 1A: Estimated date that the earliest vaccine effect would be detectable; Phase 1B: aged 65+, non-healthcare residential settings, and frontline essential workers; Phase 1C: aged 16-64 with underlying medical conditions and all other essential workers; Phase 2: remaining Chicagoans aged 16+). **(b)** Histogram of vaccination levels. **(c)** Map of zip codes.

**(a)**

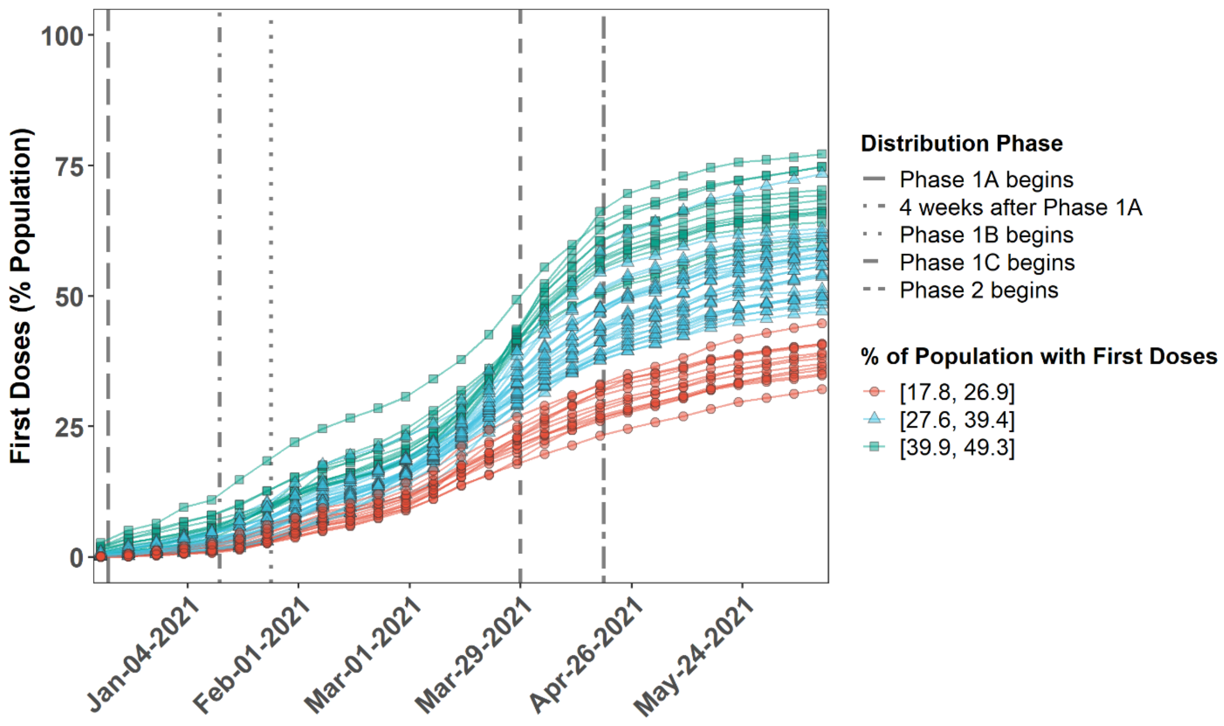

(b)

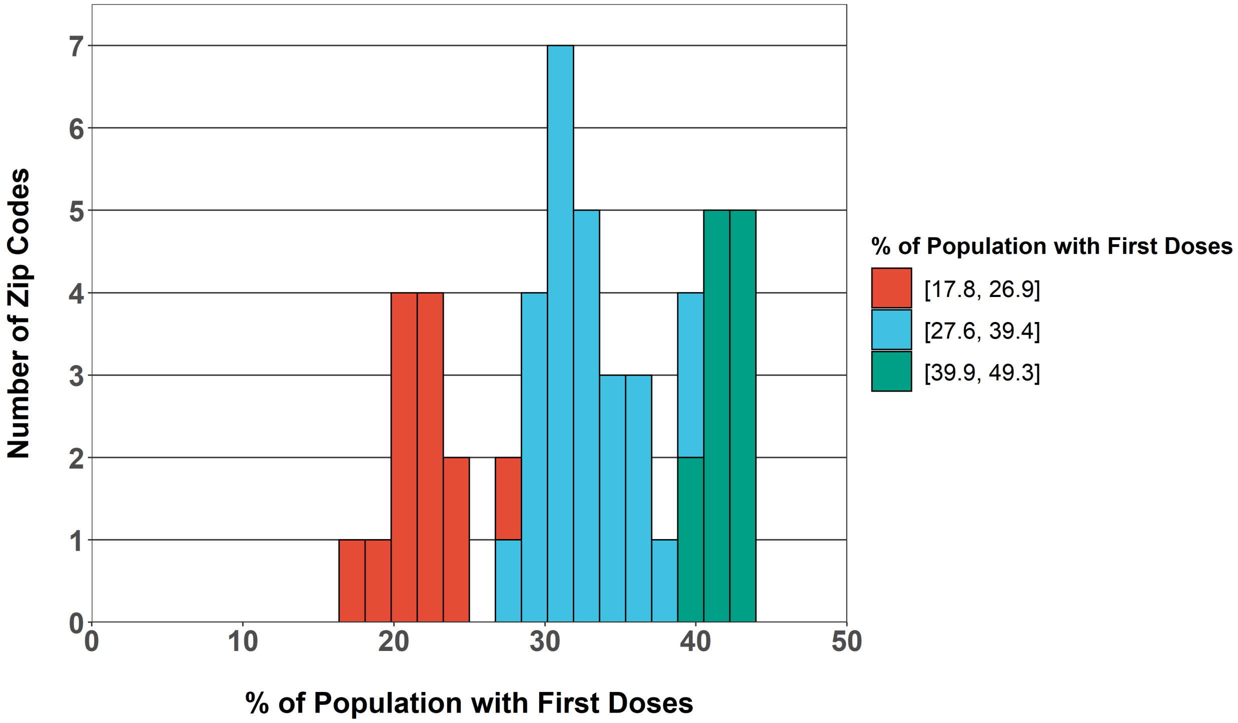
Number of Zip Codes
0
1
2
3
4
5
6
7
0
10
20
30
40
50
% of Population with First Doses
% of Population with First Doses
[17.8, 26.9]
[27.6, 39.4]
[39.9, 49.3]

(c)

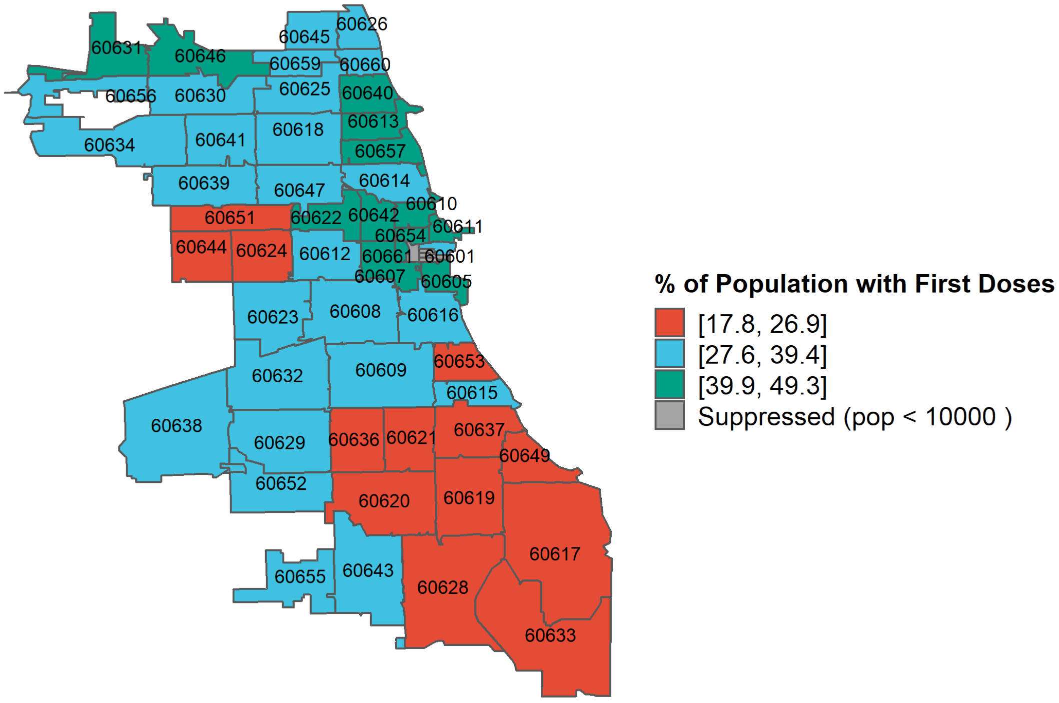
60626
60645
60631
60646
60659
60660
60656
60630
60625
60640
60613
60634
60641
60618
60657
60639
60647
60614
60610
60651
60622
60642
60611
60654
60644
60624
60612
60661
60601
60607
60605
60623
60608
60616
60632
60609
60653
60615
60638
60629
60636
60621
60637
60649
60652
60620
60619
60617
60655
60643
60628
60633
% of Population with First Doses
[17.8, 26.9]
[27.6, 39.4]
[39.9, 49.3]
Suppressed (pop < 10000 )

**Table S1: List of zip codes and groups included in analysis**

| ZIP | Vaccination Group | First Doses by 3/28/21 | % First Doses | Population |
|---|---|---|---|---|
| **60621** | [17.8, 26.9] | 4997 | 17.8 | 28018 |
| **60633** | [17.8, 26.9] | 2342 | 18.5 | 12689 |
| **60624** | [17.8, 26.9] | 6972 | 20.0 | 34892 |
| **60636** | [17.8, 26.9] | 6201 | 20.7 | 30024 |
| **60644** | [17.8, 26.9] | 9952 | 21.4 | 46591 |
| **60649** | [17.8, 26.9] | 9990 | 21.4 | 46633 |
| **60620** | [17.8, 26.9] | 15079 | 22.3 | 67711 |
| **60637** | [17.8, 26.9] | 10681 | 22.6 | 47300 |
| **60628** | [17.8, 26.9] | 14680 | 22.8 | 64254 |
| **60617** | [17.8, 26.9] | 19256 | 23.0 | 83553 |
| **60619** | [17.8, 26.9] | 14958 | 24.4 | 61207 |
| **60651** | [17.8, 26.9] | 15785 | 24.9 | 63492 |
| **60653** | [17.8, 26.9] | 8920 | 26.9 | 33154 |
| **60659** | [27.6, 39.4] | 11801 | 27.6 | 42735 |
| **60656** | [27.6, 39.4] | 8159 | 28.9 | 28218 |
| **60612** | [27.6, 39.4] | 9832 | 29.1 | 33735 |
| **60638** | [27.6, 39.4] | 17111 | 29.2 | 58669 |
| **60609** | [27.6, 39.4] | 17910 | 29.4 | 60939 |
| **60623** | [27.6, 39.4] | 24570 | 30.2 | 81283 |
| **60652** | [27.6, 39.4] | 13102 | 30.2 | 43447 |
| **60629** | [27.6, 39.4] | 33889 | 30.8 | 110029 |
| **60634** | [27.6, 39.4] | 23302 | 31.0 | 75082 |
| **60641** | [27.6, 39.4] | 21887 | 31.3 | 69880 |
| **60639** | [27.6, 39.4] | 27757 | 31.5 | 88204 |
| **60645** | [27.6, 39.4] | 14973 | 31.7 | 47270 |
| **60630** | [27.6, 39.4] | 18616 | 33.0 | 56433 |
| **60643** | [27.6, 39.4] | 16161 | 33.1 | 48887 |
| **60632** | [27.6, 39.4] | 29868 | 33.2 | 89857 |
| **60616** | [27.6, 39.4] | 18036 | 33.3 | 54197 |

| | | | | |
|---|---|---|---|---|
| **60626** | [27.6, 39.4] | 16843 | 33.3 | 50544 |
| **60625** | [27.6, 39.4] | 27436 | 34.5 | 79444 |
| **60615** | [27.6, 39.4] | 14097 | 34.7 | 40590 |
| **60618** | [27.6, 39.4] | 33035 | 34.8 | 94907 |
| **60655** | [27.6, 39.4] | 10126 | 35.4 | 28569 |
| **60647** | [27.6, 39.4] | 32117 | 36.6 | 87633 |
| **60660** | [27.6, 39.4] | 16461 | 37.0 | 44498 |
| **60601** | [27.6, 39.4] | 5801 | 38.5 | 15083 |
| **60608** | [27.6, 39.4] | 31581 | 39.4 | 80059 |
| **60614** | [27.6, 39.4] | 28380 | 39.4 | 71954 |
| **60610** | [39.9 ,49.3] | 16176 | 39.9 | 40548 |
| **60646** | [39.9 ,49.3] | 11520 | 40.3 | 28569 |
| **60642** | [39.9 ,49.3] | 8084 | 41.0 | 19716 |
| **60640** | [39.9 ,49.3] | 28609 | 41.2 | 69363 |
| **60622** | [39.9 ,49.3] | 22072 | 41.4 | 53294 |
| **60607** | [39.9 ,49.3] | 12197 | 41.6 | 29293 |
| **60631** | [39.9 ,49.3] | 12275 | 41.6 | 29529 |
| **60605** | [39.9 ,49.3] | 12378 | 42.6 | 29060 |
| **60661** | [39.9 ,49.3] | 4426 | 42.7 | 10354 |
| **60657** | [39.9 ,49.3] | 30545 | 43.0 | 70958 |
| **60613** | [39.9 ,49.3] | 21960 | 43.3 | 50761 |
| **60654** | [39.9 ,49.3] | 8728 | 43.6 | 20022 |
| **60611** | [39.9 ,49.3] | 16396 | 49.3 | 33224 |

**Table S2: Zip codes excluded from analysis**

List of zip codes excluded from analyses. Five zip codes (60602, 60603, 60604, 60606, 60666) were excluded because they contained fewer than 10,000 residents. 60707 and 60827 were excluded because they lie mostly outside the City of Chicago limits.

| ZIP | Vaccination Group | First Doses by 3/28/21 | % First Doses | Population |
|---|---|---|---|---|
| 60602 | Outliers | 702 | 613 | 1145 |

| | | | | |
|---|---|---|---|---|
| 60603 | Outliers | 709 | 67.4 | 1052 |
| 60604 | Outliers | 457 | 55.5 | 823 |
| 60606 | Outliers | 1584 | 48.2 | 3287 |
| 60666 | Outliers | 80 | 0 | 0 |
| 60707 | Outliers | 4827 | | |
| 60827 | Outliers | 469 | | |

**Table S3: Demographics of study population**

| **Age** | **No. (%)[a]** |
|---|---|
| Age 0-17 | 561320 (21) |
| Age 18-65 | 1790772 (67) |
| Age 65+ | 334263 (12) |
| **Sex** | |
| Female | 1378658 (51) |
| Male | 1307697 (49) |
| **Race and Ethnicity** | |
| Latinx | 773938 (29) |
| Asian Non-Latinx | 175220 (7) |
| Black Non-Latinx | 783916 (29) |
| White Non-Latinx | 894555 (33) |
| Other Race Non-Latinx | 58726 (2) |
| **High School Graduate or Higher** | 1582411 (84)[b] |
| **With Health Insurance** | 2402389 (90)[c] |
| | **$ (IQR)** |
| **Median Household Income** | 52044 (51437) |

[a] No. (%) = total number across group (percentage of total group population)
[b] Calculated as percentage of group population over 25 years old
[c] Calculated as percentage of group civilian noninstitutionalized population

**Table S4: Test for parallel trends**

To test the parallel trends assumption during the decline in second wave COVID-19 deaths, we fit the following model to the zip code-level data from December 13, 2020 to March 28, 2021:

(1) $\ln(Y_i) - \ln(Population) = \beta_0 + \beta_1(Week) + \beta_2(Vaccination\_Group) +$

$$\beta_3(Week \times Vaccination\_Group)$$

Where $Week \times Vaccination\_Group$ describes the effect of $Vaccination\_Group$ on weekly slope during the decline of the second wave. An F-test for $Week \times Vaccination\_Group[17.8,\ 26.9] = Week \times Vaccination\_Group[27.6, 39.4] = 0$ with 2 degrees of freedom resulted in a Chi-squared probability of 1.825 with a p-value of 0.4015. Thus, we fail to reject the null hypothesis that there was not a significant association of $Vaccination\_Group$ on the weekly slope.

| **Predictors** | **Estimate** | **Std. Error** | **p** |
|---|---|---|---|
| (Intercept) | -10.15 | 0.13 | <0.001 |
| Week | -0.13 | 0.02 | <0.001 |
| Vaccination_Group [17.8, 26.9] | 0.44 | 0.16 | 0.007 |
| Vaccination_Group [27.6, 39.4] | 0.59 | 0.14 | <0.001 |
| Week * Vaccination_Group [17.8, 26.9] | 0.02 | 0.02 | 0.412 |
| Week * Vaccination_Group [27.6, 39.4] | -0.00 | 0.02 | 0.961 |
| Observations | 832 | | |
| $R^2$ Nagelkerke | 0.488 | | |

**Table S5: Complete Mixed-Effects Models Output**

To analyze the correlation between vaccination levels on March 28, 2021 and spring wave-specific COVID-19 mortality, we narrowed our dataset to start at December 13, 2020, the peak of deaths during the second wave of COVID-19 mortality. We fit the data to the following models:

(2) $ln(Y_i) - ln(Population) = \beta_{0i} + \beta_1(Week) + \beta_2(Wave) + \beta_3(Wave \times Vaccinations)$

(3) $ln(Y_i) - ln(Population) = \beta_{0i} + \beta_1(Week) + \beta_2(Wave) + \beta_3(Wave \times Vaccinations) + \beta_4(Recovered) + \beta_5(65+)$

Where $\beta_{0i} = \beta_0 + v_{0i}$ and is a randomly distributed intercept with random effects structure

$$v_{o_i} \sim N(0, \sigma_v^2)$$

$Week$ is a continuous variable describing the number of weeks since the start of the data, $Wave$ is a categorical variable describing whether at date was in the second or spring wave,

$Recovered$ is a continuous variable describing the percentage of zip code residents who had recovered from COVID-19 by March 28, 2021, and $65+$ is a continuous variable describing the percentage of zip code residents who are over 65 years old. $Wave \times Vaccinations$ describes the effect of a 10% increase in vaccinations by March 28, 2021 on the number of deaths per week in the spring wave.

| | Unadjusted | | | Adjusted | | |
|---|---|---|---|---|---|---|
| **Predictors** | **Estimate** | **Std. Error** | **p** | **Estimate** | **Std. Error** | **p** |
| (Intercept) | -9.87 | 0.08 | <0.001 | -11.56 | 0.28 | <0.001 |
| Week | -0.11 | 0.01 | <0.001 | -0.11 | 0.01 | <0.001 |
| Wave | 2.41 | 0.28 | <0.001 | 2.39 | 0.27 | <0.001 |
| Wave×Vaccinations | -0.50 | 0.09 | <0.001 | -0.49 | 0.08 | <0.001 |
| Recovered | | | | 0.85 | 0.19 | <0.001 |
| 65+ | | | | 0.74 | 0.13 | <0.001 |
| **Random Effects** | | | | | | |
| $\sigma^2$ | 0.75 | | | 0.75 | | |
| $\tau_{00}$ | 0.18 $_{zip}$ | | | 0.07 $_{zip}$ | | |
| ICC | 0.19 | | | 0.08 | | |
| N | 52 $_{zip}$ | | | 52 $_{zip}$ | | |
| Observations | 1404 | | | 1404 | | |
| Marginal $R^2$ / Conditional $R^2$ | 0.295 / 0.430 | | | 0.396 / 0.446 | | |

**Table S6: Sensitivity Analyses**

We ran Model 3 using the percentage of residents fully vaccinated by March 28, 2021, rather than the percentage of residents with at least one dose by March 28, 2021. A 1 unit increase of $Wave \times Percentage\_Complete$ corresponds to a 10% increase in fully vaccinated residents. As a second sensitivity analysis, we ran Model 3 using the categorical variable $Week \times Vaccination\_Group$ rather than continuous variable $Wave \times Vaccinations$.

| | Percentage_Complete | | | Vaccination_Group | | |
|---|---|---|---|---|---|---|
| **Predictors** | **Estimate** | **IRR (CI)** | **p** | **Estimate** | **IRR (CI)** | **p** |
| (Intercept) | -11.63 | 0.00 (0.00 – 0.00) | <0.001 | -11.66 | 0.00 (0.00 – 0.00) | <0.001 |
| Week | -0.11 | 0.90 (0.88 – 0.91) | <0.001 | -0.11 | 0.90 (0.88 – 0.91) | <0.001 |
| Wave | 2.24 | 9.39 (5.69 – 15.49) | <0.001 | 0.42 | 1.53 (1.03 – 2.26) | 0.034 |
| Recovered | 0.85 | 2.34 (1.61 – 3.41) | <0.001 | 0.92 | 2.52 (1.70 – 3.73) | <0.001 |

| | | | | | | |
|---|---|---|---|---|---|---|
| 65+ | 0.80 | 2.22 (1.73 – 2.84) | <0.001 | 0.76 | 2.13 (1.65 – 2.76) | <0.001 |
| Wave×Percentage_Complete | -0.80 | 0.45 (0.34 – 0.59) | <0.001 | | | |
| Wave×Vaccination_Group [17.8, 26.9] | | | | 0.92 | 2.52 (1.70 – 3.73) | <0.001 |
| Wave×Vaccination_Group [27.6, 39.4] | | | | 0.33 | 1.39 (0.95 – 2.03) | 0.089 |
| **Random Effects** | | | | | | |
| $\sigma^2$ | 0.75 | | | 0.75 | | |
| $\tau_{00\ zip}$ | 0.07 | | | 0.08 | | |
| ICC | 0.08 | | | 0.09 | | |
| $N_{zip}$ | 52 | | | 52 | | |
| Observations | 1404 | | | 1404 | | |
| Marginal $R^2$ / Conditional $R^2$ | 0.386 / 0.437 | | | 0.381 / 0.437 | | |
